# SARS-CoV-2 hyperimmune globulin for severely immunocompromised patients with COVID-19: a randomised, controlled, double-blind, phase 3 trial

**DOI:** 10.1101/2022.04.04.22273314

**Authors:** Sammy Huygens, Quincy Hofsink, Inger S Nijhof, Abraham Goorhuis, Arnon P Kater, Peter AW te Boekhorst, Francis Swaneveld, Věra MJ Novotný, Susanne Bogers, Matthijs RA Welkers, Grigorios Papageorgiou, Bart J Rijnders, Jarom Heijmans

**Affiliations:** Erasmus University Medical Center, Department of Internal Medicine, Section of Infectious Diseases and Department of Medical Microbiology and Infectious Diseases, Doctor Molewaterplein 40, Rotterdam, The Netherlands; Amsterdam UMC location University of Amsterdam, Department of Hematology, Meibergdreef 9, Amsterdam, The Netherlands; Amsterdam UMC location Vrije Universiteit Amsterdam, Department of Hematology, Meibergdreef 9, Amsterdam, The Netherlands; St. Antonius Hospital, Department of Internal Medicine-Hematology, Koekoekslaan 1, Nieuwegein, The Netherlands; Amsterdam UMC location University of Amsterdam, Department of Infectious Diseases, Meibergdreef 9, Amsterdam, The Netherlands; Erasmus University Medical Center, Department of Hematology, Doctor Molewaterplein 40, Rotterdam, The Netherlands; Sanquin Blood Supply, Unit of Transfusion Medicine, Plesmanlaan 125, Amsterdam, The Netherlands; Erasmus University Medical Center, Department of Viroscience, Doctor Molewaterplein 40, Rotterdam, The Netherlands; Amsterdam UMC location University of Amsterdam, Department of Medical Microbiology & Infection Prevention, Meibergdreef 9, Amsterdam, The Netherlands; Erasmus University Medical Center, Department of Biostatistics, Doctor Molewaterplein 40, Rotterdam, The Netherlands; Amsterdam UMC location University of Amsterdam, Department of Internal Medicine, Meibergdreef 9, Amsterdam, The Netherlands

## Abstract

**Background:** Severely immunocompromised patients are at risk for severe COVID-19. Benefit from convalescent plasma in these patients is suggested but data from randomised trials are lacking. The aim of this study is to determine efficacy of SARS-CoV-2 hyperimmune globulin (“COVIG”) in treatment of severely immunocompromised, hospitalised COVID-19 patients.

**Methods:** In this randomised, controlled, double-blind, multicentre, phase 3 trial, severely immunocompromised patients who were hospitalised with symptomatic COVID-19 were randomly assigned (1:1) to receive 15 grams of COVIG or 15 grams of intravenous immunoglobulin without SARS-CoV-2 antibodies (IVIG, control). Patients included were solid organ transplant patients with three drugs from different immunosuppressive classes or patient with disease or treatment severely affecting B-cell function. Patients that required mechanical ventilation or high flow nasal oxygen were excluded. All investigators, research staff, and participants were masked to group allocation. The primary endpoint was occurrence of severe COVID-19 evaluated up until day 28 after treatment, defined as the need for mechanical ventilation, high-flow nasal oxygen, readmission for COVID-19 after hospital discharge or lack of clinical improvement on day seven or later. This trial is registered with Netherlands Trial Register (NL9436).

**Findings:** From April, 2021, to July, 2021, 18 participants were enrolled at three sites in the Netherlands; 18 patients were analysed. Recruitment was halted prematurely when casirivimab/imdevimab became the recommended therapy in the Dutch COVID-19 treatment guideline for seronegative, hospitalised COVID-19 patients. Median age was 58 years and all but two were negative for SARS-CoV-2 spike IgG at baseline. Severe COVID-19 was observed in two out of ten (20%) patients treated with COVIG compared to seven of eight (88%) in the IVIG control group (*p* = 0·015, Fisher’s exact test).

**Interpretation:** COVIG reduced the incidence of severe COVID-19 in severely immunocompromised patients, hospitalised with COVID-19. COVIG may be a valuable treatment in this patient group and can be used when no monoclonal antibody therapies are available.

**Funding:** The Netherlands Organisation for Health Research and Development, Sanquin Blood Supply Foundation.

## Introduction

Severely immunocompromised patients, such as organ transplant recipients and patients with hematologic malignancies, are at risk for a severe course of COVID-19 with higher rates of hospital admission, more severe pulmonary disease and increased mortality rates.^1,2^ The increased burden of disease in these patients may at least be partly due to an impaired ability to generate antibodies upon infection or after vaccination.^3–6^ In contrast to immunocompetent patients, of which the majority has already seroconverted upon hospital admission for COVID-19, immunocompromised patients are often still SARS-CoV-2 seronegative at the time of admission.^7^

Although an unprecedented number of trials have been completed and have contributed to our understanding of antibody therapy with convalescent plasma and highly potent monoclonal antibodies for hospitalised COVID-19 patients, immunocompromised patients have been grossly underrepresented in these trials. Furthermore, as far as we know, there has not been a single randomised controlled intervention trial that focused on immunocompromised patients specifically, despite the fact that this population is likely to benefit most from such interventions.

When a new viral disease arises, convalescent plasma is usually the most rapidly available antibody-based therapy. In high risk elderly patients, a reduced rate of hospitalisation was observed when convalescent plasma was administered within 72 hours after disease onset and before hospital admission was required.^8^ In contrast, in hospitalised patients, convalescent plasma did not improve outcome, even in patients who were seronegative at the time of treatment.^9–16^ Together, these results have led the world health organization (WHO) to advise against treatment with convalescent plasma in hospitalised COVID-19 patients. Regarding the use of SARS-CoV-2 hyperimmune globulin (“COVIG”) among hospitalised COVID-19 patients, only three trials were performed. One phase I/II trial randomised 50 hospitalised COVID-19 patients to COVIG or standard of care and demonstrated a lower mortality in the COVIG arm (25% in COVIG arm versus 60% in control group), however, this benefit could not be confirmed in two phase III trials.^17–19^ Highly potent virus neutralizing monoclonal or combined monoclonal antibodies yield supraphysiological levels of neutralizing antibodies unachievable with plasma-derived products. The landmark trial by the UK RECOVERY group evaluating treatment with a combination of casirivimab and imdevimab in hospitalised patients showed reduced mortality, but only in the predefined subgroup of seronegative patients.^20^

In severely immunocompromised patients, uncontrolled observational data suggest a benefit of convalescent plasma for the treatment of COVID-19, but this has not been confirmed in randomised trials.^21–31^ We set out to examine effects of COVIG in a double-blind, controlled, randomised fashion.

## Methods

### Study design and participants

The COVID-Compromise trial was a randomised, controlled, double-blind, multicentre, phase 3 trial to evaluate the effects of COVIG in severely immunocompromised patients hospitalised with COVID-19. The trial was intended for enrolment in ten hospitals in the Netherlands, but owing to the early termination of the trial, patients were enrolled in only 3 hospitals. The study complied with IRB/EC procedures, the Declaration of Helsinki, ICH Good Clinical Practices guideline, the EU directive Good Clinical Practice (2001-20-EG), and local regulations governing the conduct of clinical studies. The protocol was approved by the medical ethics committees of all participating centres, and written informed consent was obtained from all patients.

Adult patients who were severely immunocompromised as defined in the study protocol (see supplementary data file 1) and hospitalised with a PCR-confirmed symptomatic SARS-CoV-2 infection were eligible for inclusion within 72 hours after admission. Patients that had received prior treatment with convalescent plasma or IVIG with neutralizing SARS-CoV-2 antibodies, patients that had documented hypersensitivity to IVIG, or patients that required respiratory support with endotracheal intubation or high flow nasal oxygen were excluded.

### Randomisation and masking

Eligible and consenting patients were randomly assigned in a 1:1 fashion to receive either 15 grams COVIG or 15 grams of an identical looking product without SARS-CoV-2 antibodies (“IVIG”). COVIG contained a neutralising titre of 900 IU/ml (VNT50) against SARS-CoV-2 wild-type variant.^32^ Randomisation was performed by computer, stratified according to the type of the immunocompromised state. Both COVIG and IVIG were produced by Prothya, the Netherlands, and labelled similarly as Nanogam®. All investigators, research staff, and participants were blinded to the allocated treatment until day 28, but unblinding was possible before day 28 when the primary endpoint was reached.

### Clinical and biochemical monitoring

Baseline data were collected using a web-based case report form, including demographics, medical history, vital parameters and respiratory support. At baseline and following treatment SARS-CoV-2 antibodies were determined and nasal swabs were collected for the quantification of viral *RNA*. Serum SARS-CoV-2 antibody measurements were performed using LIAISON® SARS-CoV-2 TrimericS IgG assay (DiaSorin). Positivity was defined as spike IgG >33·8 BAU/ml.

### Outcomes

The primary endpoint of this study was occurrence of severe COVID-19 evaluated up until day 28 after treatment. Severe COVID-19 was defined as any of the following: (1) an indication for high-flow nasal oxygen, (2) need for mechanical ventilation, (3) lack of clinical improvement after at least seven days of observation after treatment or (4) readmission for PCR-confirmed COVID-19 within 28 days. Lack of clinical improvement was defined as no decrease in oxygen requirement on day seven or any day thereafter compared to the highest level of oxygen requirement, or, in patients that did not require oxygen, persistence of fever not due to alternative causes or in patients without fever no clinical improvement as evaluated by the blinded treating physician.

Secondary endpoints included occurrence of severe COVID-19 in the subgroup of patients that had no SARS-CoV-2 antibodies upon inclusion, duration of hospitalisation, 28-day mortality, the four individual endpoints that compose the primary endpoint, rate of viral decay and safety.

### Early trial termination

The trial was initiated in April, 2021, when monoclonal antibody therapy was not available as standard of care for COVID-19. In mid-June, 2021, monoclonal antibodies casirivimab/imdevimab became recommended for seronegative, hospitalised COVID-19 patients by the Dutch COVID-19 treatment guideline, based on the results of the RECOVERY trial.^20^ Consequently, on July 29, 2021, after discussion with the data safety monitoring board, the study team decided to stop enrolment as it was no longer deemed ethically acceptable to withhold casirivimab/imdevimab therapy for patients in the trial.

### Statistical analysis

We estimated that this high-risk patient group had a 70% chance of reaching the primary endpoint of severe COVID-19, and hypothesized a reduction to 30% with COVIG treatment. With a power of 90% and a two-sided alpha of 5% and a single pre-planned efficacy interim analysis, a sample size of 86 participants was required. Due to premature termination of the study, analysis was performed after enrolment of 21% of the target population. Analysis was performed in the modified intention-to-treat population. Continuous variables were described as medians with interquartile ranges (IQR). Categorical variables were described as proportions. In the primary endpoint analysis, proportions in both treatment groups were compared by a Fisher’s exact test, given the small number of observations.

Repeated measurements of viral decay were analysed using a mixed-effects model that included a linear effect of time, an interaction between time and treatment group and random intercepts. The model was adjusted for age, whether a patient was vaccinated, and duration of symptoms at inclusion. Based on this model the average rate of change of viral decay per treatment group was quantified via the regression coefficient of the interaction term between time and treatment group.

## Results

From April, 2021, to July, 2021, a total of 37 patients was screened of which 18 were enrolled at three sites in the Netherlands. Ten patients were randomly allocated to receive SARS-CoV-2 hyperimmune globulin (“COVIG”) and eight to conventional IVIG without SARS-CoV-2 antibodies (Figure 1).

**Figure 1:**
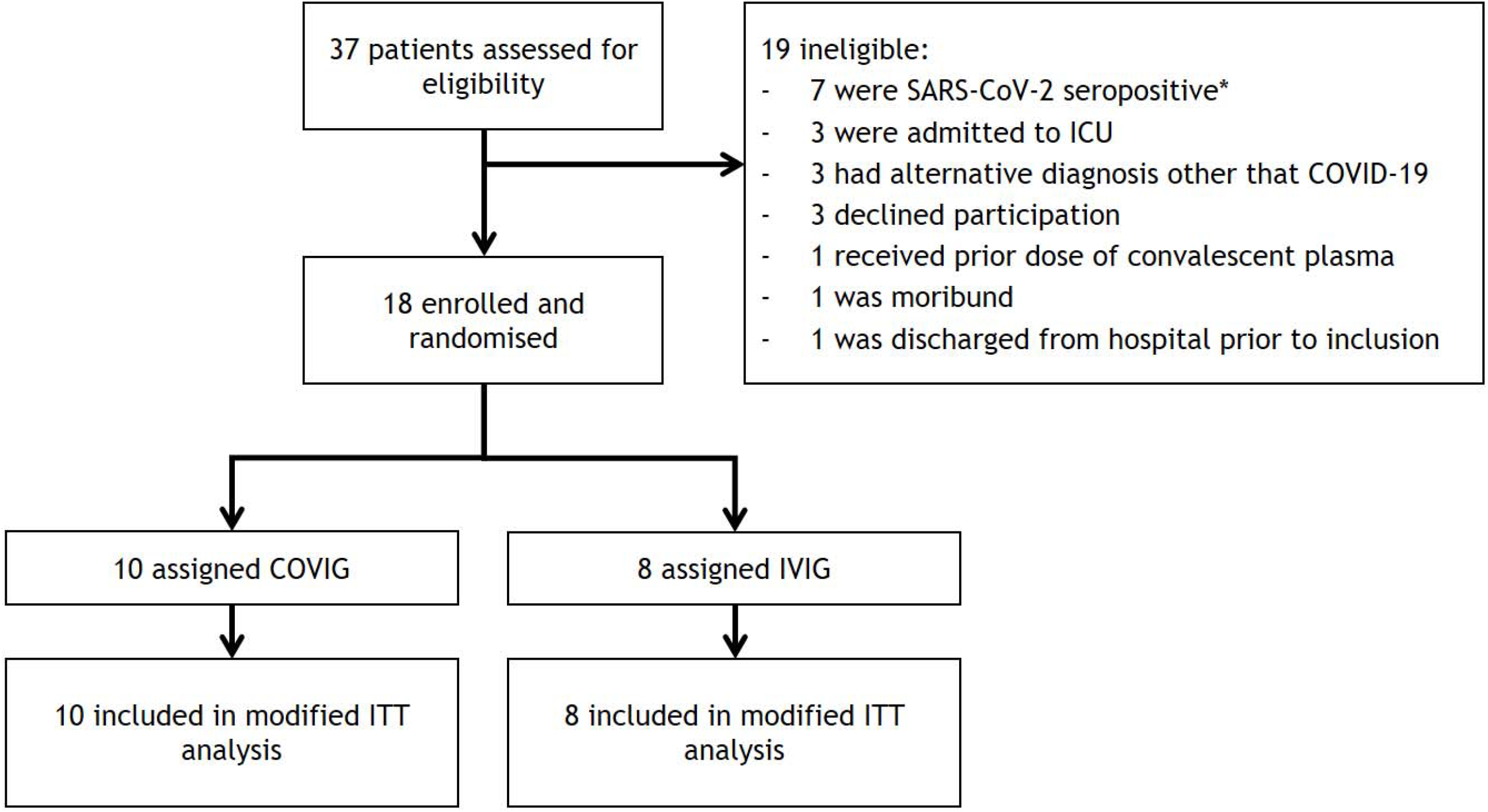
Trial profile. *organ transplant recipients requiring immunosuppressive medication from two pharmacological classes could be included when seronegative only

At baseline, median age of the patients was 58 years (IQR 35 – 66) and symptoms had been present for nine days (IQR 7 – 21). The median Charlson Comorbidity Index was 3 (IQR 2 – 5) and oxygen supplementation was 3 L/min (IQR 0 – 5). Two patients (11%) had SARS-CoV-2 IgG antibodies upon inclusion (Table 1).

**Table 1.** Demographic and baseline characteristics.

|  | **COVIG group**<br>**(n = 10)** | **IVIG group**<br>**(n = 8)** |
| --- | --- | --- |
| Age (yr) – median (IQR) | 48 (27 – 61) | 63 (41 – 72) |
| Male gender – no. (%) | 6 (60) | 4 (50) |
| BMI<sup>1</sup> (kg/m<sup>2</sup>) – median (IQR) | 26 (24 – 32) | 29 (24 – 33) |
| Charlson Comorbidity Index – median (IQR) | 2 (2 – 4) | 5 (2 – 7) |
| Cause of immunocompromised state – no. (%) |  |  |
| Immunosuppression for solid organ transplant | 4 (40) | 5 (63) |
| Recent use of anti-B cell therapy for malignancy | 3 (30) | 3 (38) |
| Immunosuppression for auto-immune disease | 1 (10) | 0 (0) |
| Congenital hypogammaglobulinemia | 1 (10) | 0 (0) |
| Acquired hypogammaglobulinemia | 1 (10) | 0 (0) |
| Prior vaccination against SARS-CoV-2 |  |  |
| One or more | 5 (50) | 8 (100) |
| Full schedule<sup>2</sup> | 5 (50) | 6 (75) |
| Number of days since symptom onset – median (IQR) | 10 (8 – 22) | 8 (6 – 26) |
| Number of days since admission to hospital – median (IQR) | 2 (1 – 4) | 1 (1 – 2) |
| Corticosteroid therapy for COVID-19 at baseline<sup>3</sup> – no. (%) | 10 (100) | 4 (50) |
| Tocilizumab therapy for COVID-19 at baseline – no. (%) | 1 (10) | 0 (0) |
| Respiratory support |  |  |
| No oxygen supplementation – no. (%) | 1 (10) | 4 (50) |
| Simple oxygen supplementation<sup>4</sup> – no. (%) | 9 (90) | 4 (50) |
| Required oxygen dose (L/min) – median (IQR) | 3 (1 – 8) | 1 (0 – 4) |
| SARS-CoV-2 variant |  |  |
| Wild-type – no. (%) | 1 (10) | 1 (13) |
| Alpha – no. (%) | 5 (50) | 5 (63) |
| Unknown – no. (%) | 4 (40) | 2 (25) |
| Presence SARS-CoV-2 IgG – no. (%) | 2 (20) | 0 (0) |
| B-cell count (per μL) – median (IQR)<sup>5</sup> | 12 (0 – 144) | 20 (0 – 40) |
| CD4 T-cell count<sup>6</sup> | 111 (65 – 257) | 147 (53 – 298) |
| CD8 T-cell count<sup>6</sup> | 140 (71 – 227) | 180 (140 – 245) |

<sup>1</sup> BMI was not available for 1 patient in both groups.
<sup>2</sup> Patient was considered as fully vaccinated 7 days after a 2<sup>nd</sup> vaccination with Moderna or BioNTech/Pfizer COVID-19 vaccine, 14 days after Janssen COVID-19 vaccine or after one Moderna or BioNTech/Pfizer COVID-19 vaccine in combination with prior SARS-CoV-2 infection.
<sup>3</sup> All patients that required supplemental oxygen received corticosteroid therapy per standard of care.
<sup>4</sup> Oxygen supplementation via nasal cannula.
<sup>5</sup> Counts were not available for 3 patients in COVIG group.

|  |  |  |
| --- | --- | --- |
| NK-cell count<sup>6</sup> | 60 (41 – 90) | 90 (50 – 176) |

The primary endpoint analysis included data from all 18 patients (figure 2 and table 2). Severe COVID-19 occurred in two (20%) patients in the COVIG arm compared to seven (88%) patients in the IVIG (p = 0·015). Among all 16 seronegative patients, 13% developed severe COVID-19 in the COVIG arm compared to 88% in the IVIG arm. Among the two seropositive patients, one patient developed severe COVID-19 and both were treated with COVIG. All separate parameters that composed the primary endpoint were more frequent in the IVIG arm compared to the COVIG arm. The median duration of hospital stay was nine days in the COVIG group (IQR 4 – 15) and nine days in the IVIG group (IQR 4 – 17). The protocol allowed unblinding and cross-over to the intervention arm when a patient reached the primary endpoint. Therefore, five of eight patients randomised to IVIG eventually also received COVIG or convalescent plasma as therapy.

**Table 2.** Primary and secondary outcomes^1^.

|  | **COVIG group**<br>**(n = 10)** | **IVIG group**<br>**(n = 8)** | **RR (95% CI)** | **Fisher's exact test (p)** |
| --- | --- | --- | --- | --- |
| Severe course of COVID-19 | 2 (20) | 7 (88) | 0.23 (0.06 – 0.81) | 0.015 |
| Severe course of COVID-19 in seronegative patients | 1/8 (13) | 7/8 (88) | 0.14 (0.02 – 0.91) |  |
| Mortality at 28 days | 0 (0) | 3 (38) |  |  |
| Median duration of hospitalization<sup>2</sup> | 9 (4 to 15) | 9 (4 to 17) |  |  |
| Indication for adjunctive ventilator support | 2 (20) | 5 (63) |  |  |
| Admission to an intensive care unit due to respiratory insufficiency | 1 (10) | 3 (38) |  |  |
| Lack of clinical improvement at day 7 or any day thereafter | 0 (0) | 3 (38) |  |  |
| Readmission for COVID-19 | 0 (0) | 2 (25) |  |  |

<sup>1</sup> Data are n (%), median (IQR) or n/N (%).
<sup>2</sup> Analyses exclude those who died during hospitalization.

**Figure 2:**
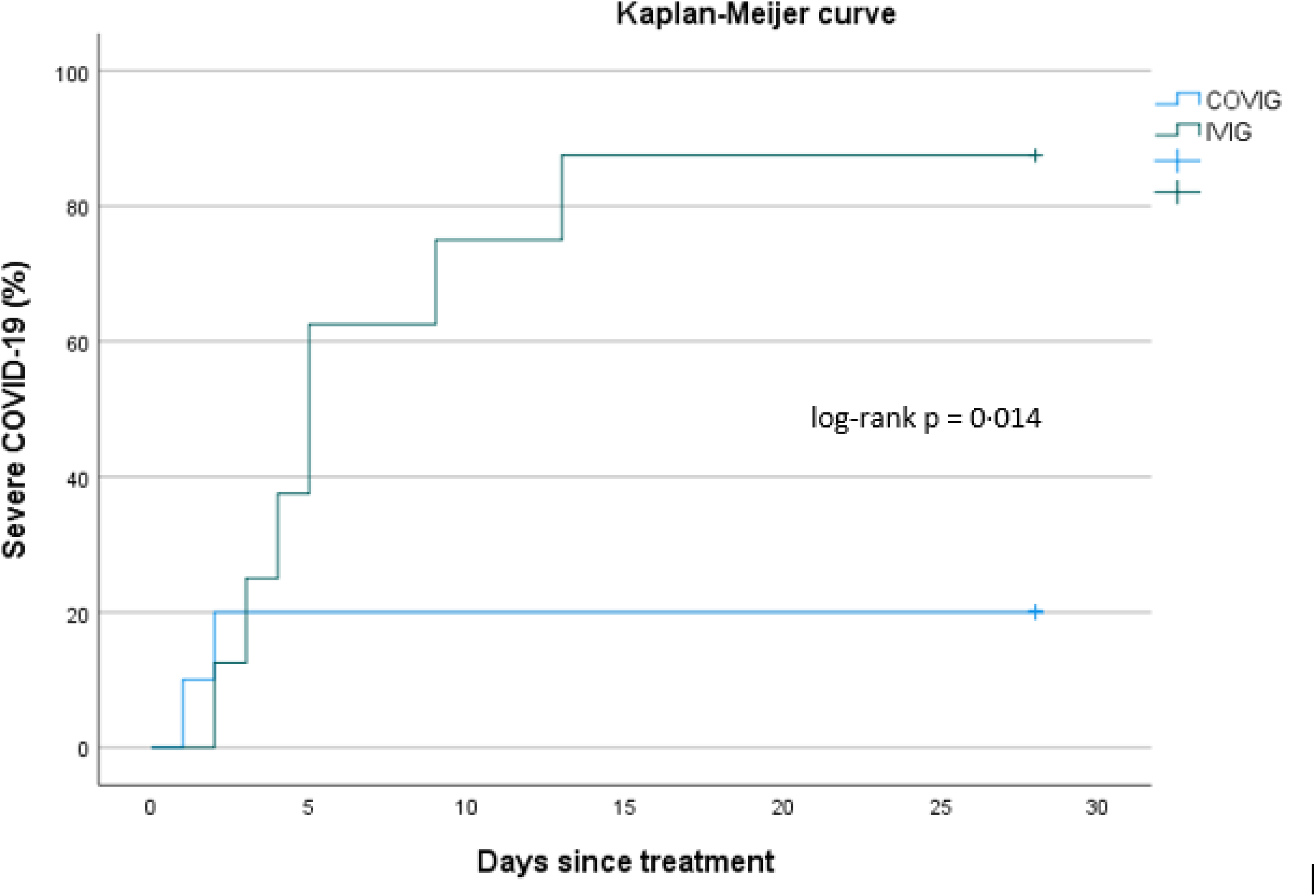
Occurrence of severe COVID-19 per group. Severe COVID-19 was defined as any of the following: (1) an indication for high-flow nasal oxygen, (2) need for mechanical ventilation, (3) lack of clinical improvement after at least 7 days of observation after treatment, or (4) readmission for PCR-confirmed COVID-19 within 28 days.

Of all 18 patients, nine patients (50%) had a severe adverse event (SAE) of which eight were related to COVID-19. There were three deaths due to COVID-19, all in the IVIG group. One patient was readmitted for treatment of a community-acquired bacterial pneumonia and after antibiotic treatment, he fully recovered within ten days. No infusion related serious adverse events were observed.

No difference in the viral decay over time was observed between the two groups (supplementary figure 1).

## Discussion

In this randomised, controlled, double-blind trial we aimed to determine the effect of SARS-CoV-2 hyperimmune globulin (“COVIG”) as a treatment for COVID-19 in severely immunocompromised patients. Treatment with COVIG reduced the incidence of severe COVID-19 from 88 to 20% (p = 0·015). No patient died in the COVIG arm while three of eight patients in the control arm died.

The study was terminated prematurely when casirivimab/imdevimab became available as standard of care for hospitalised, seronegative, COVID-19 patients in the Netherlands, resulting in a small sample size. However, the absolute treatment effect we observed was higher than we had anticipated in the study protocol. These effects may be explained by higher age and comorbidity index in the control arm resulting in more severe COVID-19 in this group. However, in the COVIG arm, patients had a higher baseline oxygen requirement.

A number of trials has evaluated efficacy of convalescent plasma or COVIG for hospitalised COVID-19 patients and results have so far not been in favour of these interventions. However, very few immunocompromised patients were included.^7–14,16,17,19,33^ The only information we have in severely immunocompromised patients, is from case series where effects of convalescent plasma were evaluated in an uncontrolled setting. Therefore, and despite the small sample size, the results of our randomised, double-blind trial are an important addition to the current evidence in favour of COVIG for severely immunocompromised patients with COVID-19. This treatment is of particular importance in a context where monoclonal antibody therapy is unavailable or when new variants of concern emerge that are resistant against most or all registered monoclonal antibodies.

The surprisingly strong effect of COVIG on disease course in severely immunocompromised patients, compared to absent effects of equipotent doses of convalescent plasma in immunocompetent patient may pinpoint that therapeutic dose and moment of antibody-based treatment is less critical in the patient population that have hampered antibody production.

We found no difference in viral decay between the two groups, suggesting that viral load in the upper respiratory tract does not correlate well with clinical outcome at day 28. A similar observation was made in an outpatient, placebo-controlled trial with the antiviral drug remdesivir, where treatment resulted in 87% protection from hospitalisation.^34^ Moreover, animal studies have shown that higher doses of neutralizing monoclonal antibodies are needed to decrease the viral load in the upper airways compared to the lower respiratory tract.^35,36^

In this trial, all participants were infected with the SARS-CoV-2 wild-type (Wuhan-1) or alpha variant and the COVIG that was utilized was generated by collecting convalescent plasma of patients who had recovered after infection with the SARS-CoV-2 wild-type (Wuhan-1). Currently, the B.1.1.529 variant has become the dominant variant and in many parts of the world the BA.2 variant is becoming dominant. The B.1.1.529 variant is resistant to most monoclonal antibodies currently available, with sotrovimab as an exception.^37,38^ However, several research groups reported a strongly reduced activity of sotrovimab to the BA.2 variant.^38,39^ This clearly illustrates that convalescent plasma or COVIG may continue to have a role to play in the treatment of severely immunocompromised patients with COVID-19. However, it is crucial that this plasma or COVIG is able to neutralize the variant that the patient is infected with. This should be possible when plasma donors with very high antibody titres are selected (e.g. shortly after booster vaccination).^40^

In summary, in severely immunocompromised patients, COVIG may reduce the risk for severe COVID-19 and can be used when no monoclonal antibody therapies are available.

## Supporting information

Study protocol

## Data Availability

All data produced in the present work are contained in the manuscript.

