## Supplementary material for "SARS-CoV-2 hyperimmune globulin for severely immunocompromised patients with COVID-19: a randomised, controlled, double-blind, phase 3 trial": Study protocol

### Convalescent Antibody-Mediated Treatment of COVID-19 Infections in Patients with B-cell dysfunction, a Randomized Trial

***COVID-Compromise Study***

**FINAL**

| | |
|---|---|
| **Date:** | **30 March2021** |
| **Principal Investigator:** | **Dr. J. Heijmans** |
| **Sponsor:** | **AmsterdamUMC** |
| **EudraCT number:** | **2020-006075-15** |

**PROTOCOL TITLE** 'Antibody medicated treatment of COVID-19 infections in immunocompromised patients'

| | |
|---|---|
| **Protocol ID** | **COVID Compromise** |
| **Short title** | **COVID compromise study** |
| **EudraCT number** | **2020-006075-15** |
| **Version** | **3** |
| **Date** | **30 March 2021** |
| **Coordinating investigator** | **Dr. Jarom Heijmans**<br>**Amsterdam UMC Location AMC**<br>**Meibergdreef 9**<br>**1105 AZ Amsterdam**<br>**The Netherlands**<br>**** |
| **Principal Investigator**<br><br><br>**Co-Principal Investigator** | **Dr. Jarom Heijmans**<br>****<br>**Tel: +31 20 7329255**<br>**Prof. Dr. AP Kater**<br>****<br>**tel: +31 20 7329255**<br>**Dr. B.J.A. Rijnders**<br>****<br>**tel: Tel: +31 10 7033510** |
| **Sponsor** | **Amsterdam UMC location AMC**<br>**Meibergdreef 9**<br>**1105 AZ Amsterdam**<br>**The Netherlands** |
| **Subsidising party** | **Sanquin Blood Supply BV** |
| **Study Statistician** | **Greg Papageorgiou**<br>**Erasmus MC** |
| **Independent expert** | **Prof. Dr. H. van Laarhoven**<br>**Amsterdam UMC, Location VUmc**<br>**Head of Medical Oncology Department**<br>**** |
| **Laboratory sites** | **Sanquin** |

| | |
|---|---|
| | Department of Experimental Immunohematology, Sanquin Research<br>Plesmanlaan 125<br>1066 CX Amsterdam<br>The Netherlands<br>Tel: +31 20 512 3333<br>Fax: +31 20 512 3332<br>Contactperson: Prof. dr. E. v.d. Schoot |
| Pharmacy<br><br><br><br><br><br>Bloodbank | Kenniscentrum Amsterdam UMC, location AMC<br>Meibergdreef 9<br>1105 AZ Amsterdam<br>The Netherlands<br>Contact person: Dr. E.M. Kemper<br><br>Sanquin Blood Supply<br>Plesmanlaan 125<br>1066 CX Amsterdam<br>The Netherlands<br>Tel: +31 20 512 3333<br>Fax: +31 20 512 3332<br>Contact person: Drs. F.H. Swaneveld |
| Writing Committee | Prof. Dr. M.J. Kersten, Amsterdam UMC, location AMC<br>Dr. V.M.J. Novotny, Sanquin<br>Dr. F. Swaneveld, Sanquin<br>Dr. A. Goorhuis, Amsterdam UMC, location AMC<br>Dr. B.J.A. Rijnders, ErasmusMC<br>Dr. P.A.W. te Boekhorst, ErasmusMC<br>Dr. I.S. Nijhof, Amsterdam UMC, location VUmc<br>Dr. A.H.W. Bruns, UMC Utrecht |

**PROTOCOL SIGNATURE SHEET**

| Name | Signature | Date |
|---|---|---|
| **Sponsor or legal representative:**<br>**Prof. Dr. A.P. Kater**<br>**Head of Department of Hematology**<br>**Amsterdam UMC location AMC** | | |
| **Coordinating Investigator:**<br>**Dr. Jarom Heijmans, Hematologist**<br>**Amsterdam UMC location AMC** | | |

#### SITE PRINCIPAL INVESTIGATOR SIGNATURE PAGE

| **Local Site Name:** | **Signature** | **Date** |
|---|---|---|
| **Name of Site Principal Investigator:** | | |

By my signature, I agree to personally supervise the conduct of this study in my affiliation and to ensure its conduct in compliance with the protocol, informed consent, IRB/EC procedures, the Declaration of Helsinki, ICH Good Clinical Practices guideline, the EU directive Good Clinical Practice (2001-20-EG), and local regulations governing the conduct of clinical studies.

#### SUMMARY OF CHANGES

The following substantive changes were made as part of this amendment:

| Change | Rationale |
|---|---|
| Paragraph 8.2: Stratification of randomization has changed. Randomization is stratified according to group of immunocompromised origin | When stratifying for age, gender and cause of immunocompromised origin, too many strata result. We have chosen to do multivariate analyses to correct for age and gender and stratify for ICP cause only. |
| Paragraph 6.4, 7.4 and 8.2: Blinding of Nanogam plus changed. This will be performed using VTGM (“voor toediening gereed maken”) stickers with specific identification numbers for the study and the patient instead of the GMP annex 13. | According to the trial pharmacist, using VTGM stickers is sufficient and much more time efficient, since the medication will be administered clinically and does not concern novel medication. This is in line with IGJ guidelines. These stickers will cover the only distinguishing part of the Nanogam vs Nanogam plus flask which is the batch number. |
| Paragraph 8.2: Unblinding can also be done by the local pharmacy | Since the study is multicenter, it is important that emergency unblinding is feasible in all centers at all times. Not all centers are familiar with automatic emergency unblinding in CASTOR and since the pharmacy is not blinded, they may consult with the local pharmacist in emergency settings. |
| Paragraph 9.4 and 13.6: Biological material will be stored for 5 years instead of 15 years | Longer storage is not required. |

#### LIST OF ABBREVIATIONS AND RELEVANT DEFINITIONS

| | |
|---|---|
| ABR | General Assessment and Registration form (ABR form), the application form that is required for submission to the accredited Ethics Committee; in Dutch: Algemeen Beoordelings- en Registratieformulier (ABR-formulier) |
| AE | Adverse Event |
| AR | Adverse Reaction |
| CA | Competent Authority |
| CCMO | Central Committee on Research Involving Human Subjects; in Dutch: Centrale Commissie Mensgebonden Onderzoek |
| CLL | Chronic lymphocytic leukemia |
| CV | Curriculum Vitae |
| DSMB | Data Safety Monitoring Board |
| EU | European Union |
| EudraCT | European drug regulatory affairs Clinical Trials |
| GCP | Good Clinical Practice |
| GDPR | General Data Protection Regulation; in Dutch: Algemene Verordening Gegevensbescherming (AVG) |
| HCL | Hairy Cell Leukemia |
| HSCT | Hematopoietic Stem Cell Transplantation |
| IB | Investigator's Brochure |
| IC | Informed Consent |
| ICP | Immunocompromised Patient |
| IMP | Investigational Medicinal Product |
| IMPD | Investigational Medicinal Product Dossier |
| IVIG | Intravenous Immunoglobulins |
| METC | Medical research ethics committee (MREC); in Dutch: medisch-ethische toetsingscommissie (METC) |
| PLL | Prolyphocytic Leukemia |
| (S)AE | (Serious) Adverse Event |
| SPC | Summary of Product Characteristics; in Dutch: officiële productinformatie IB1-tekst |
| Sponsor | The sponsor is the party that commissions the organisation or performance of the research, for example a pharmaceutical |

| | |
|---|---|
| | **company, academic hospital, scientific organisation or investigator. A party that provides funding for a study but does not commission it is not regarded as the sponsor, but referred to as a subsidising party.** |
| **SUSAR** | **Suspected Unexpected Serious Adverse Reaction** |
| **TACO** | **Transfusion Associated Cardiac Overload** |
| **TRALI** | **Transfusion Associated Lung Injury** |
| **UAVG** | **Dutch Act on Implementation of the General Data Protection Regulation; in Dutch: Uitvoeringswet AVG** |
| **WMO** | **Medical Research Involving Human Subjects Act; in Dutch: Wet Medisch-wetenschappelijk Onderzoek met Mensen** |

#### SYNOPSIS

**Rationale:**

A more severe course of COVID-19 infections has been reported in immunocompromised patients (ICPs), such as those suffering from innate immune disorders, (haematological) malignancies, or patients who are treated with immunosuppressive medication. The common denominator among these patients is an impaired B-cell or T-cell function, or depletion, and these patients are unable to generate sufficient neutralizing antiviral antibody titres or their production is delayed. To date, no specific therapeutic strategies have convincingly been shown effective in this patient population. Treatment with neutralizing antiviral antibodies from convalescent patients has been tested in the general population. In this population, one study has shown promising results when it is administered in the very first stages of disease, but not in hospitalized patients. This discrepancy is thought to result from the redundant effect of exogenous antibodies in these patients, in whom high titres of neutralizing antibodies are typically present at hospital admission. In those who are still antibody negative at admission, all but few start producing antibodies within days after admission as well. Since immunocompromised patients may not develop neutralizing antibodies, exogenously provided antibodies could be a functional adjuvant therapy of COVID-19, also in later stages of disease (e.g. upon hospitalization), but no trials to data have addressed this question.

**Objective**:

<u>Primary Objectives:</u>

To generate proof that convalescent antibody-mediated treatment of ICPs, who suffer from COVID-19 disease can protect ICPs from a more severe course of disease.

<u>Secondary objective:</u>

- To evaluate the impact of convalescent antibody-mediated treatment on duration of hospitalization in the ICP population.
- To evaluate the impact of convalescent antibody-mediated treatment on mortality in the ICP population.
- To evaluate the impact of convalescent antibody-mediated treatment on ICU admission in the ICP population.
- To evaluate the impact of convalescent antibody-mediated treatment on symptom duration in the ICP population.
- To evaluate the impact of convalescent antibody-mediated treatment on viral clearance in the ICP population.

**Study design:**

The trial is designed as a cohort specific, randomized, controlled, comparative, phase 2/3 study.

**Study population:**

This study includes ICPs that are hospitalized for COVID-19. ICP is here defined as patients in whom the underlying disease itself or immunosuppressive treatment resulted in a probable B-cell dysfunction or depletion, resulting in a reduced potential to generate anti-SARS-CoV-2 antibodies.

Key Inclusion criteria:

- Patient is ≥ 18 years of age, diagnosed with COVID-19 based on a positive PCR or antigen test.
- Hospitalized.

AND one of immunocompromised conditions/treatments below

**B-cell inhibition related ICP**

- Use of anti-CD19 or -CD20 directed antibody therapy in 6 months prior to inclusion.
- Previous or current treatment with drugs that significantly impair B cell function (e.g. ibrutinib, venetoclax, acalabrutinib, idelalisib etc) within 6 months prior to inclusion

**Other immunosuppression/treatment related ICP**

- Patients treated with bendamustine, purine analogues or anti-thymocyte globulin within 6 months prior to inclusion.
- Solid organ transplant patients that are taking systemic immunosuppressive drugs from at least three pharmacological classes.

**Cellular therapy related ICP**

- Allogeneic hematopoietic stem cell transplant (HSCT) in 12 months prior to inclusion.
- HSCT for which systemic therapy against graft-versus-host-disease is used.
- Recipient of CAR-T cells < 2 years prior to inclusion.

**Disease related ICP**

- Chronic B-cell leukemia´s: CLL, HCL, PLL, multiple myeloma, Waldenströms macroglobulinemia

**Congenital ICP**

- Congenital disorder resulting in severe B-cell dysfunction or depletion requiring immunoglobulin suppletion (e.g. agammaglobulinemia).

<u>Key Exclusion criteria:</u>

- Has previously participated in this study.
- Has previously received convalescent plasma with high level neutralizing anti-SARS-CoV-2 antibodies (either in other study or in compassionate use program).
- Known hypersensitivity to treatment with immunoglobulins (WHO grade 3-4).
- Patient who has reached endpoint already at admission (direct adjunctive oxygen therapy in the form of high-flow nasal oxygen (HFNO), mechanical ventilation or ICU admission for other reason).

**Target number of patients**:
86 patients (43 receive study treatment, 43 receive control treatment).

**Intervention**:
After randomization, one group (43 patients) receives treatment with Nanogam containing high titre neutralizing anti-SARS-CoV-2 antibodies (further referred to as *Nanogam plus*). The other group (43 patients) receives control treatment with Nanogam without high titre anti SARS-CoV-2 antibodies as control for Nanogam plus. Treatment is in a double blinded fashion.

**Main study parameters/endpoints:**
The endpoint of this study is a severe course of disease, evaluated up until day 28 after randomization:

- Start of adjunctive ventilator support (HFNO, mechanical ventilation) or an indication to do so but due to due to previously agreed treatment restrictions HFNO or mechanical ventilation is not initiated.
- Admission to an intensive care unit for progression of respiratory insufficiency.
- No clinical improvement on day 7 after randomization or any day thereafter after first day of treatment (based on oxygen use in patients that require oxygen and based on clinical disease burden (including fever) in patients that require no oxygen).
- Readmission for COVID-19.

<u>Secondary endpoints</u>
To evaluate the impact of convalescent antibody-mediated treatment in the ICP population on:

- Severity of COVID-19 disease in patients that have no anti SARS-CoV-2 antibodies upon inclusion (‘per protocol’ analysis).
- Duration of hospitalization.

- 28-day overall mortality
- The four individual endpoints that compose the primary endpoint.
- Time to complete recovery from COVID-19 related symptoms.
- Rate of viral decay
- Development of long-term persistent neutralizing antibodies against SARS-CoV-2
- T-cell immunity as measured by in vitro specific T cell response to COVID-19 tetrameric antigens.

**Nature and extent of the burden and risks associated with participation:**
Infusion of Nanogam plus or control (Nanogam); Viral PCR on day 1, 3, 7, 10, 14 and weekly until discharge, then on months 2, 3, 6, 12. Blood withdrawal weekly until discharge (mostly covered during standard of care blood withdrawal) and monthly
Monthly follow-up measurement of anti-SARS-CoV-2 antibodies in serum until 3 months after discharge. Monthly follow-up measurement of SARS-CoV-2 virus by PCR until 3 months after discharge.
The risks of Nanogam infusion are small and widely known. These consist mainly of infusion reactions or volume overload.
Benefits of this study may include a shorter disease course and a decrease in mortality.

#### 1. INTRODUCTION AND RATIONALE

The COVID-19 pandemic has caused significant morbidity and mortality all over the world. COVID-19, the clinical syndrome resulting from infection with the SARS-CoV-2 virus, has a highly variable course that differs from a largely non-symptomatic or mildly symptomatic upper respiratory tract disease to a severe viral pneumonia with high levels of inflammation and thrombosis. A number of factors that carry increased risk for severe disease or death have been described, of which high age is most important (Banerjee, Pasea et al. 2020, Ioannidis 2020).

Immunocompromised patients (ICP), especially those that suffer from reduced B- or T- cell function or depletion, have an increased risk of a severe course of numerous viral diseases (Chemaly, Ghosh et al. 2006, Aksoy, Harputluoglu et al. 2007). Diagnosis of (hematologic) malignancy or treatment with immunosuppressive drugs are marked as risk factors for severe COVID-19 disease with an associated mortality rate of up to 30% (Jee, Foote et al. 2020, Parra-Bracamonte, Lopez-Villalobos et al. 2020). To date, a number of treatment modalities for COVID-19 have been proposed, of which treatment with steroids in combination with supportive care measures remains the mainstay (guideline 2020). A number of antibody mediated therapies have been proposed, including convalescent plasma, hyperimmune gammaglobulins and monoclonal (or oligoclonal) antibodies. To date, none of these therapies have been proven successful in mitigating the severity of COVID-19 disease in larger cohorts of COVID-19 infected patients (covid19treatmentguidelines.nih.gov). An important explanation lies in the rapid development of host neutralizing antibodies in almost 80% of severe COVID-19 patients (Gharbharan a 2020). However, no trials have been performed on antibody mediated treatment modalities to prevent or treat COVID-19 infections in ICPs, who may not be able to develop these neutralizing antibodies.

#### 2. OBJECTIVES

Primary Objective:
To generate proof that convalescent antibody-mediated treatment of ICPs, who suffer from COVID-19 disease can protect ICPs from a more severe course of disease.

Secondary Objective:

- To evaluate the impact of convalescent antibody-mediated treatment on duration of hospitalization in the ICP population.
- To evaluate the impact of convalescent antibody-mediated treatment on mortality in the ICP population.
- To evaluate the impact of convalescent antibody-mediated treatment on ICU admission in the ICP population.
- To evaluate the impact of convalescent antibody-mediated treatment on symptom duration in the ICP population.
- To evaluate the impact of convalescent antibody-mediated treatment on viral clearance in the ICP population.

#### 3. STUDY DESIGN

The study will be performed as a nationwide multicenter 1:1 randomized trial to compare efficacy of convalescent antibody mediated therapy to control. Treatment is with Nanogam containing high titer neutralizing anti-SARS-CoV-2 antibodies (further referred to as *Nanogam plus*) for which the control treatment is Nanogam from batches that contain low or no levels of neutralizing anti-SARS-CoV-2 antibodies.

Upon reaching the endpoints of the study, treatment of patients will be unblinded. The treating physician may consider compassionate use convalescent plasma in selected individuals (there is no compassionate use program for Nanogam plus currently) when study treatment was with control agent. Further analyses of anti-SARS-CoV-2 antibody titers and PCR for SARS-CoV-2 will be performed as standard of care.

Randomization will be done via a web-based system (CASTOR). In this system, the investigator can randomize the patient while the result of the randomization will not be disclosed to the investigator but to a designated unblinded person from the pharmacy.

### 4. STUDY POPULATION

#### 4.1 Population

Eligible patients are adults 18 years of age or older who have severely impaired B-cell function or have severe B-cell depletion (see inclusion criteria) and who are hospitalized because of COVID-19, regardless of the duration of symptoms.

Patients will be informed on the possibility of a referral for study participation by the treating physician of the patient.

The treating physician will inform the study team when a patient has given permission to do so. The patient may be transferred to a study centre for participation. In the hospital, written informed consent will be obtained.
For patients that are hospitalized for support or treatment of COVID-19 related complaints, inclusion should be within 72 hours since admission.
For patients that are hospitalized for other causes but develop COVID-19 related symptoms, inclusion should be within 72 hours since positive PCR for SARS-CoV-2.

#### 4.2 Inclusion criteria

- Patient is ≥ 18 years of age, diagnosed with COVID-19 based on a positive PCR or antigen test in combination with COVID-19 related symptoms (e.g. Fever, hypoxia, gastrointestinal symptoms).
- Hospitalized.

AND one of immunocompromised conditions/treatments below

**B-cell inhibition related ICP**

- Use of anti-CD19 or -CD20 directed antibody therapy in 6 months prior to inclusion.
- Previous or current treatment with drugs that significantly impair B cell function (e.g. ibrutinib, venetoclax, acalabrutinib, idelalisib etc) within 6 months prior to inclusion

**Other immunosuppression/treatment related ICP**

- Patients treated with bendamustine, purine analogues or anti-thymocyte globulin within 6 months prior to inclusion.
- Solid organ transplant patients that are taking systemic immunosuppressive drugs from at least three pharmacological classes. Or from at least two classes in combination with negative anti-SARS-CoV-2 antibodies ≤ 96 hours prior to inclusion.

**Cellular therapy related ICP**

- Allogeneic hematopoietic stem cell transplant (HSCT) in 12 months prior to inclusion.
- HSCT for which systemic therapy against graft-versus-host-disease is used.
- Recipient of CAR-T cells < 2 years prior to inclusion.

**Disease related ICP**

- Chronic B-cell leukemia´s: CLL, HCL, PLL, multiple myeloma, Waldenströms macroglobulinemia.

**Congenital ICP**

- Congenital disorder resulting in severe B-cell dysfunction or depletion requiring immunoglobulin suppletion (e.g. agammaglobulinemia).

##### 4.3 Exclusion criteria

A potential subject who meets any of the following criteria will be excluded from participation in this study:

- Patient or legal representative is unable to provide written informed consent
- Life expectancy of < 28 days in the opinion of the treating physician
- Has previously participated in this study
- Has previously received convalescent plasma with high level neutralizing anti-SARS-CoV-2 antibodies (either in other study or in compassionate use program).
- Known IgA deficiency (defined as absence of IgA and possibility of anti-IgA antibodies), patients with disease related reduced levels of IgA (e.g. in myeloma or lymphoma) may be included in the study.
- Known previous grade 3 or 4 hypersensitivity reactions to treatment with immunoglobulins.
- Patient who has reached endpoint already at admission (direct adjunctive oxygen therapy in the form of high-flow nasal oxygen (HFNO), mechanical ventilation or ICU admission for other reason).

#### 5. TREATMENT OF SUBJECTS

##### 5.1 Investigational product/treatment

<u>Use of Nanogam Plus</u>

We will administer Nanogam plus. The procedure for generation of Nanogam plus is identical to production of Nanogam, with exception that the donors for the Nanogam plus batch are selected on presence of high titer neutralizing anti-SARS-CoV-2 antibodies.

##### 5.2 Use of co-intervention (if applicable)

As co-medication for Nanogam plus or control, antihistaminic drugs will be given to patients who have had prior reactions to immunoglobulin concentrates: clemastin 2mg orally or intravenously prior to infusion of Nanogam plus.

In patients in whom hypoxia results from COVID-19, dexamethasone 6mg is given for a maximal duration of 10 days, alternative medications that are given per standard of care for COVID-19 related disease may be given to all patients.

##### 5.3 Escape medication (if applicable)

In case of the occurrence of an allergic reaction, the attending physician may prescribe standard treatment for allergic reactions consisting of intramuscular treatment with adrenalin, antihistaminic drugs and/or steroids according to local protocol.

#### 6. INVESTIGATIONAL PRODUCT

##### 6.1 Name and description of investigational product(s)

Nanogam plus:

Nanogam plus is generated according to established procedure by which Nanogam is generated with exception that for production, patients that have high titer neutralizing antibodies are selected.

##### 6.2 Summary of findings from non-clinical studies

For product information or findings from non-clinical studies we refer to the Smpc of the product (see Annex # 5)

##### 6.3 Dosages, dosage modifications and method of administration

The dose of Nanogam plus shall be calculated based on the amount of neutralizing antibodies present in the batch that will be used at that moment. This dose will match the amount of neutralizing antibodies that is present in the convalescent plasma pool (e.g. 600 ml 1:160 neutralizing titre). In the first batch of Nanogam plus, a neutralizing titre of 1:160 is present, therefore 600 ml will be administered. In the second batch of Nanogam plus, a fourfold higher level of neutralizing antibodies results in a dose of 150 ml that will be administered. Nanogam plus will be administered intravenously on one day per standard of care for administration of Nanogam as described in the label.

#### 6.4 Preparation and labelling of Investigational Medicinal Product

Preparation and labelling of the investigational product is done using VTGM (“voor toediening gereed maken”) stickers with specific identification numbers for the study.

#### 6.5 Drug accountability

Stocks of Nanogam plus will be delivered to the pharmacy of the participating hospital. A Confirmation of Receipt form should be sent to SBB. When a patient is eligible for entering the study, the investigator should order Nanogam plus as study medication at the pharmacy of the hospital. Study medication not used for the study should be returned to SBB or destroyed in consultation with SBB.

The investigator, or a pharmacist or other appropriate individual who is designated by the investigator, should maintain records of the product’s delivery to the trial site, the inventory at the site, the use by each patient, and the return to the sponsor or alternative disposition of unused product(s). These records should include dates, quantities, batch/serial numbers, expiration dates (if applicable), and the unique code numbers assigned to the investigational product(s) and trial patients (if applicable). Investigators should maintain records that document adequately that the patients were provided the doses specified by the protocol and reconcile all investigational product(s) received from the sponsor. The investigator should also collect and count remaining medication and empty boxes of medication to check that the patient has taken the assigned dose.

#### 6.6 Study drug return and destruction

Partially used investigational medicinal product should not be redispensed to either the same or another patient after it has been returned.
The trial site should destroy used or partially used study drug containers after drug accountability records have been completed. Destruction should be documented.

### 7. NON-INVESTIGATIONAL PRODUCT

#### 7.1 Name and description of non-investigational product(s)

As comparative product for the use of Nanogam plus, Nanogam will be administered.

#### 7.2 Summary of findings from non-clinical studies

For product information or findings from non-clinical studies we refer to the smpc of the product (see Annex # 5)

#### 7.3 Dosages, dosage modifications and method of administration

Patients will receive a dose of Nanogam (similar amount of grams of immunoglobulins) equal to the calculated volume of Nanogam plus (see section 6.3). Nanogam is infused per standard of care, described in the label.

#### 7.4 Preparation and labelling of Non Investigational Medicinal Product

Preparation and labelling of the investigational product is done using VTGM (“voor toediening gereed maken”) stickers with specific identification numbers for the study.

#### 7.5 Drug accountability

Stocks of Nanogam will be delivered to the pharmacy of the participating hospital. A Confirmation of Receipt form should be sent to SPP. When a patient is eligible for entering the study, the investigator should order Nanogam as study medication at the pharmacy of the hospital. Study medication not used for the study should be returned to SBB or destroyed in consultation with SBB.

The investigator, or a pharmacist or other appropriate individual who is designated by the investigator, should maintain records of the product’s delivery to the trial site, the inventory at the site, the use by each patient, and the return to the sponsor or alternative disposition of unused product(s). These records should include dates, quantities, batch/serial numbers, expiration dates (if applicable), and the unique code numbers assigned to the investigational product(s) and trial patients (if applicable). Investigators should maintain records that document adequately that the patients were provided the doses specified by the protocol and reconcile all investigational product(s) received from the sponsor. The investigator should also collect and count remaining medication and empty boxes of medication to check that the patient has taken the assigned dose.

##### 7.6 Drug return and destruction

All Nanogam plus that is not used for this study will be returned to SBB.

#### 8. METHODS

##### 8.1 Study parameters/endpoints

###### 8.1.1 Main study parameter/endpoint

The endpoint of this study is a severe course of disease, evaluated up until day 28 after randomization:

- Start of adjunctive ventilator support (HFNO, mechanical ventilation) or an indication to do so but due to due to previously agreed treatment restrictions HFNO or mechanical ventilation is not initiated.
- Admission to an intensive care unit for progression of respiratory insufficiency.
- No clinical improvement on day 7 after randomization or any day thereafter after first day of treatment.
    - In patients in need of oxygen therapy during first 7 days after randomization this is defined as:
        - Use of a non-rebreather mask with flow of ≥ 10 liters for a period of ≥ 6 hours in order to reach a saturation of ≥ 93% during rest or mild effort*.
        - No decrease in the required oxygen supplementation (of at least 2 liters/min) compared to the highest required oxygen supplementation in the first seven days since hospital admission (defined as highest required amount of oxygen supplementation for ≥ 6 hours to reach a saturation of ≥ 93% during rest or mild effort)*.
    - In patients not in need of oxygen therapy at any time during first seven days this is defined as lack of clinical improvement compared to the highest disease burden in the first seven days according to the treating physician.
    - Persistent or recurrent fever (defined as ≥ 38.5 C) on day 7 or any day thereafter after first day of treatment without any alternative focus of infection or fever.
- Readmission for COVID-19.

* Low saturation should be attributed to COVID-19 and not to apparent alternative causes such as obstructive sleep apnoea or hearth failure.

###### 8.1.2 Secondary study parameters/endpoints (if applicable)

- Severity of COVID-19 disease in patients that have no anti SARS-CoV-2 antibodies upon inclusion (‘per protocol’ analysis).
- Duration of hospitalization.
- 28 days overall mortality.
- The four individual endpoints that compose the primary endpoint.
- Time to complete recovery from COVID-19 related symptoms.
- Rate of viral decay
- Development of long-term persistent neutralizing antibodies against SARS-COV-2
- T-cell immunity as measured by in vitro specific T cell response to COVID-19 tetrameric antigens.

###### 8.1.3 Other study parameters (if applicable)

Age and comorbidity (diabetes, obesity, cardiovascular risk) are known risk factors that segregate patients that have a high risk for a severe disease course.

##### 8.2 Randomisation, blinding and treatment allocation

Eligible patients should be registered before start of treatment. Patients need to be registered in the electronic case record form. In principle, the registration and randomization procedure can be modified in line with institutional specifics, but should be similar to the following logistics:

All eligibility criteria will be checked with a checklist before the patient is included and randomized. Patients will be randomized with the use of an online randomization system (Castor) using blocks of up to 12 in size . Randomization is stratified according to group of immunocompromised origin (e.g. specific B cell inhibition, cellular therapy, immunosuppressive treatment, congenital). Each patient will be given a unique patient study number (a sequence number by order of enrolment in the trial). Patient study number will be given immediately by the online registration database and confirmed by email. The result of the randomization will not be disclosed to the investigator but to a designated unblinded person from the transfusion lab in the hospital. The responsible physician will order the study medication under the received study number.
Blinding of Nanogam plus will be performed using VTGM (“voor toediening gereed maken”) stickers with specific identification numbers for the study and the patient. These stickers will cover the only distinguishing part of the Nanogam vs Nanogam plus flask which is the batch number.

Unblinding procedures

While the safety of patients should always take priority, maintenance of blinding is crucial to the integrity of the double-blind part of this trial. The blind for a specific patient should only be broken when information about the patient's protocol treatment is considered necessary to manage Serious Adverse Events (emergency unblinding). Unblinding procedures should preferably be initiated only after consultation of the principal investigator or his/her representative. Emergency unblinding can be done by the site using the Emergency Unblinding form in the eCRF database. After completing and submitting the eCRF form, CASTOR will show the treatment the patient was assigned to. The AmsterdamUMC will automatically receive an email that the patient has been unblinded. After unblinding, the patient goes off protocol treatment. Alternatively, after filling out the emergency unblinding form in CASTOR, unblinding can be performed after contact with the local pharmacy.

Unblinding per protocol will be performed upon reaching the primary endpoints (e.g. requirement for adjunctive ventilation, ICU admission, no improvement on day 7 after inclusion), where unblinding based on the lack of improvement will always be discussed with the study monitor. All patients are unblinded at day 28, since the primary endpoint cannot be altered at that point.

#### 9. Study procedures

##### 9.1 Time of clinical evaluations

- Pre-screening
- Screening and baseline (day 1)
- During hospital admission: blood withdrawal according to standard care protocol of participating center.
- Daily clinical evaluation and evaluation of fever and required supplemented oxygen therapy (endpoint), evaluation of WHO ordinal scale for clinical improvement (appendix 1).
- Virological evaluation twice weekly in first two weeks, and once weekly thereafter until discharge. Once monthly until 3 months after inclusion.
- Immunological evaluation weekly until discharge and once monthly thereafter until 3 months after inclusion.

Virological and immunological examinations are partly standard of care in the study cohort, since even when COVID-19 related symptoms have abated, patients may remain viremic and may therefore be advised to quarantine themselves not based on symptomatology, but on viral parameters.

Antibody titer measurement, one 6 ml serum tube day 1, 7, 14, 21, 28, month 2, 3. Timepoint 21 may be cancelled after patient discharge.

Nasopharyngeal swab for real-time quantitative PCR on day 1, 3, 7, 10, 14, 21, 28, month 2, month 3. Timepoint 21 may be cancelled after patient discharge. When no viral clearance has been shown, patients and their physicians will be advised to continue measuring viral presence in the nasopharyngeal cavity every 2 to 4 weeks.

Immune response evaluation; four 9mL natrium heparine tubes, one 6mL EDTA tube and two 6mL serum tubes, on day 1, 7, 14, 28 (both cohorts).

#### 9.2 Schudule of Assessments

Required investigations at entry, during treatment and during follow up:

| | SOC or NOC | Prescreening with referring physician (*) | Screenin g/ Day 1 | D2 | D3 | D7 | D10 | D14 | D21 | D28 | M2 | M3 | M6 | M12 |
|---|---|---|---|---|---|---|---|---|---|---|---|---|---|---|
| Eligibility check (*) | NOC | x | x | | | | | | | | | | | |
| Informed consent | NOC | | x | | | | | | | | | | | |
| COVID PCR (**) | SOC | X | X(&) | | x | x | x | x | x | x | x | x | x | x |
| Antibody test (e.g. Wantai) | SOC | | X (&) | | | x | | x | x | x | x | x | x | x |
| Blood tests | SOC | | X (&) | x | | x | | x | | x | | x | | x |
| Nanogam plus or Nanogam infusion | NOC | | x | x | | | | | | | | | | |
| Transcutaneous O2 saturation without administration of supplemental oxygen*** | SOC | | x | x | x | x | | x | | x | x | x | | x |
| Registration of baseline characteristics and comorbidities | NOC | | x | | | | | | | | | | | |
| Telephone contact | SOC | | | | | x | | x | | x | x | x | x | x |

SOC: standard of care; NOC no standard of care.

(&) this test will be repeated when performed in a different center than where inclusion/follow up will be performed or when performed > 48 hours previously.

(*) During a telephone call with the referring physician, the in- and exclusion criteria are checked.

(**) upon clearance of COVID, no further PCR will be performed

(***) this assay will not be performed after discharge from hospital.

#### 9.3 Specification of required investigations

Please note that not all examinations that are mentioned here will be requested on the case report form (CRF). Examinations that are essential to monitor the patients' condition and patients' safety are also mentioned here, although they may not be essential for the study endpoints.

**Medical history**

Standard medical history, including:

- How was the patient referred for the study
- Sex and age
- Ethnicity
- Socio-economic status
- Body mass index (BMI) at inclusion into the study
- Date of first day of symptoms of SARS-CoV-2 infection
- Underlying medical illnesses at the time of first day of SARS-CoV-2 RT-PCR positive
- Charlson Comorbidity Index
- Clinical Frailty Scale for patients aged 70 or older
- C-reactive protein (CRP)
- SARS-CoV-2 RT-PCR cycle-time (*Ct)*-value
- Data and vaccine type of SARS-CoV-2 vaccinatino (if applicable)

**Physical examination at baseline**

- Transcutaneous O2 saturation without administration of supplemental oxygen
- Respiratory rate, blood pressure, pulse, temperature

**Laboratory testing**

- Antibody testing at t=0: Presence of anti-SARS-CoV-2 receptor binding domain (RBD)-protein and anti-SARS-CoV-2 nucleocapsid protein antibodies will be tested in a highly sensitive bridging assay (Wantai test or equivalent test). In the positive samples, the antibodies will be specified and quantified in IgG, IgA and IgM ELISA's against RBD and Nucleocapsid protein
- Nasopharynx swab for SARS-CoV-2 RT-PCR testing if > 48 hours prior to inclusion. This sample will be stored for later analysis of SARS-CoV-2 genotype.
- Level of total IgG, IgA and IgM.
- Hemogram including leukocyte level and differential

- Percentage of T-cells, (split into CD4+, CD8+ and double negative fractions), percentage of B-cells, percentage of NK-cells.
- CRP.

**Clinical follow-up**

- Fever and required supplemental oxygen use (numbers of liters of oxygen to maintain SaO2 ≥ 93% during rest or mild effort)
- COVID-19 related symptoms (improvement from baseline) and fever will be obtained by telephone on day 8, 15, 22, 29
- Any other anti-SARS-CoV-2 treatments that were given.
- Assessment of WHO ordinal scale for clinical improvement on day 7 and after completion of steroid treatment.

**Laboratory and pulmonary follow-up**

- Antibody testing on day 7, 14, 21, 28. Month 2, 3, 6, 12 (will be performed in a central lab for blinding puposes).
- SARS-CoV-2 RT-PCR Ct-values on days 3, 7, 10, 14, 21, 28. Month 2, 3, 6, 12.
- Weekly laboratory evaluation on COVID-19 ward according to treating physician minimally including CRP, hemogram and renal function.

#### 9.4 Storage for future studies (biobank)

In addition to these investigations, all patients will be asked for informed consent to store biological material for future studies. Material for future investigations will be shipped to the central laboratory of the Amsterdam UMC, location AMC. More details can be found in the study lab manual. All materials are anonymized and stored for a maximum of 5 years after end of study, after which the samples will be destroyed.

#### 9.5 Withdrawal of individual subjects

Subjects can leave the study at any time for any reason if they wish to do so without any consequences. The investigator can decide to withdraw a subject from the study for urgent medical reasons.

##### 9.5.1 Specific criteria for withdrawal

Potentially life-threatening reaction during Nanogam infusion

#### 9.6 Replacement of individual subjects after withdrawal

Patients who are withdrawn from treatment for other reasons than death will be followed for disease status as described. if they consent, SAE information will be collected as described in section 10.2.2.

#### 9.7 Follow-up of subjects withdrawn from treatment

Patients who are withdrawn from treatment for other reasons than death will be followed as described in section 9.2 for follow up. SAE information will be collected as described in section 10.2.2.

.

#### 9.8 Premature termination of the study

The sponsor may decide to terminate the study prematurely based on the following criteria:

1) There is evidence of unacceptable risks for study patients (i.e. safety issue);
2) There is ample evidence of efficacy
3) There is reason to conclude that continuation of the study cannot serve a scientific purpose

The sponsor will promptly notify all concerned investigators, the data safety monitoring board (DSMB), the ethics committee(s) and the regulatory authorities of the decision to terminate the study. The sponsor will provide information regarding the timelines of study termination and instructions regarding treatment and data collection of enrolled patients.

### 10. SAFETY REPORTING

#### 10.1 Temporary halt for reasons of subject safety

In accordance to section 10, subsection 4 of the WMO, the sponsor will suspend the study if there is sufficient ground that continuation of the study will jeopardise subject health or safety. The sponsor will notify the accredited METC without undue delay of a temporary halt including the reason for such an action. The study will be suspended pending a further positive decision by the accredited METC. The investigator will take care that all subjects are kept informed.

#### 10.2 AEs, SAEs and SUSARs

##### 10.2.1 Adverse events (AEs)

Adverse events are defined as any undesirable experience occurring to a subject during the study, whether or not considered related to [the investigational product / trial procedure/ the experimental intervention].

However, given the setting of a large outbreak, the registration of AE is not feasible and will not serve the safety of the patients. However, all AE at least possibly related to the Nanogam infusion will be registered. The side effects of Nanogam are well-known and described in the SPC. Therefore, AE that are considered unrelated to Nanogam as well as AE that are considered related will not be registered unless they have not been previously reported as per SPC.

##### 10.2.2 Serious adverse events (SAEs)

A serious adverse event is any untoward medical occurrence or effect that

- results in death;
- is life threatening (at the time of the event);
- requires hospitalisation or prolongation of existing inpatients' hospitalisation;
- results in persistent or significant disability or incapacity;
- is a congenital anomaly or birth defect; or
- any other important medical event that did not result in any of the outcomes listed above due to medical or surgical intervention, but could have been based upon appropriate judgement by the investigator.
- An elective hospital admission will not be considered as a serious adverse event.

Serious Adverse Events (SAEs) will be reported from the first administration of treatment according to protocol until 30 days following the last dose of any drug from the protocol treatment schedule or until the start of subsequent systemic therapy for the disease under study, if earlier.
Serious adverse events (including death) occurring after 30 days and/or after the start of subsequent systemic therapy should also be reported if considered at least possibly related to the investigational (medicinal) product by the investigator.

SAEs must be reported to the AmsterdamUMC, Clinical Trial Office **within 24 hours** after the event was known to the investigator, using the SAE report form provided. This initial report should contain a minimum amount of information regarding the event, associated treatment and patient identification, as described in the detail in the instructions for the SAE report form. Complete detailed information should be provided in a follow-up report within a further 2 business days, if necessary.

The sponsor will report the SAEs through the web portal *ToetsingOnline* to the accredited METC that approved the protocol, within 7 days of first knowledge for SAEs that result in death or are life threatening followed by a period of maximum of 8 days to complete the initial preliminary report. All other SAEs will be reported within a period of maximum 15 days after the sponsor has first knowledge of the serious adverse events.
All transfusion related reactions – irrespective of potential causality or not- in study patients will be recorded and analysed and reported according to national blood transfusion and haemovigilance guidelines which are standard and operational in every Dutch hospital. This is done by the institutional haemovigilance employee of each hospital as they do for all blood products administered in Dutch hospitals.

##### 10.2.3 Suspected unexpected serious adverse reactions (SUSARs)

Adverse reactions are all untoward and unintended responses to an investigational product related to any dose administered.

Unexpected adverse reactions are SUSARs if the following three conditions are met:

1. the event must be serious (see chapter 9.2.2);
2. there must be a certain degree of probability that the event is a harmful and an undesirable reaction to the medicinal product under investigation, regardless of the administered dose;

3. the adverse reaction must be unexpected, that is to say, the nature and severity of the adverse reaction are not in agreement with the product information as recorded in:
   - Summary of Product Characteristics (SPC) for an authorised medicinal product;
   - Investigator's Brochure for an unauthorised medicinal product.

The sponsor will report expedited the following SUSARs through the web portal *ToetsingOnline* to the METC:

- SUSARs that have arisen in the clinical trial that was assessed by the METC;
- SUSARs that have arisen in other clinical trials of the same sponsor and with the same medicinal product, and that could have consequences for the safety of the subjects involved in the clinical trial that was assessed by the METC.

The remaining SUSARs are recorded in an overview list (line-listing) that will be submitted once every half year to the METC. This line-listing provides an overview of all SUSARs from the study medicine, accompanied by a brief report highlighting the main points of concern. The expedited reporting of SUSARs through the web portal ToetsingOnline is sufficient as notification to the competent authority.

The expedited reporting will occur not later than 15 days after the sponsor has first knowledge of the adverse reactions. For fatal or life threatening cases the term will be maximal 7 days for a preliminary report with another 8 days for completion of the report.

##### 10.3 Annual safety report

In addition to the expedited reporting of SUSARs, the sponsor will submit, once a year throughout the clinical trial, a safety report to the accredited METC, competent authority, and competent authorities of the concerned Member States.

This safety report consists of:

- a list of all suspected (unexpected or expected) serious adverse reactions, along with an aggregated summary table of all reported serious adverse reactions, ordered by organ system, per study;
- a report concerning the safety of the subjects, consisting of a complete safety analysis and an evaluation of the balance between the efficacy and the harmfulness of the medicine under investigation.

##### 10.4 Follow-up of adverse events

All AEs (as defined in section 10.2) will be followed until they have abated, or until a stable situation has been reached. Depending on the event, follow up may require additional tests or medical procedures as indicated, and/or referral to the general physician or a medical specialist.

SAEs need to be reported till end of study within the Netherlands, as defined in the protocol

##### 10.5 Data Safety Monitoring Board (DSMB)

The DSMB will advise the Principal Investigator in writing about the continuation of the trial. The DSMB will review the general progress and feasibility of the trial, the quality and completeness of the data, adverse events and safety. The DSMB will consider if there is any concern regarding the safety and well-being of trial subjects or regarding the scientific validity of the trial results. The DSMB will base its advice on the reports provided by the statistician and the Data Manager. The DSMB is free to take into consideration external information, such as the (interim) results of other trials or literature reports.

The DSMB consists of at least three members, with at least one statistician and two physicians. Details of the DSMB constitution and tasks are documented in the trial specific DSMB charter. The DSMB will receive at least the following reports:

- Interim safety listings, including listings on SAEs and SUSARs from the trial statistician or Data Manager for review
- DSUR and annual progress reports until the final analysis of the trial primary endpoint provided by the Data Manager as notification

The advice(s) of the DSMB will only be sent to the sponsor of the study. Should the sponsor decide not to fully implement the advice of the DSMB, the sponsor will send the advice to the reviewing METC, including a note to substantiate why (part of) the advice of the DSMB will not be followed.

##### 10.6 Product Complaints

A product quality complaint (PQC) is defined as any suspicion of a product defect related to manufacturing, labeling, or packaging, i.e., any dissatisfaction relative to the identity, quality, durability, or reliability of a product, including its labeling or package integrity. A PQC may have an impact on the safety and efficacy of the product. Timely, accurate, and complete reporting and analysis of PQC information from studies are crucial for the protection of subjects, investigators, and the sponsor, and are mandated by regulatory agencies worldwide.

All initial PQCs for Nanogam must be reported to Sanquin Plasma Products by the study-site personnel within 24 hours after being made aware of the event. If the defect is combined with a serious adverse event, the investigational staff must report the PQC to Sanquin according to the serious adverse

event reporting timelines within 24 hours of their knowledge of the event. A sample of the suspected product should be maintained for further investigation if requested by Sanquin. Please also inform the AmsterdamUMC of all product complaints by email. Note that product complaints in and of themselves are not AEs. If a product complaint results in an SAE, an SAE form should be completed and sent to AmsterdamUMC (see section 10.2.2).

### 11. STATISTICAL ANALYSIS

#### 11.1 Patient numbers and power considerations

The primary goal of the study is to generate proof that convalescent antibody-mediated treatment of COVID-19 disease can accelerate recovery and protects ICPs from a severe course of disease.

Due to strict inclusion criteria and strict definition of treatment failure as the primary endpoint, we assume that treatment failure will be observed in 70% in the control arm and reduced to 35% in the treatment arm. Using the O'Brien Flemming alpha-spending function to account for one interim analysis and a 1:1 randomization, 2 groups of 43 patients are required for the study to have a power of 90% and an adjusted alpha of 4.82%.

#### 11.2 Statistical analysis

All main analyses will be according the modified intention to treat principle, i.e. restricted to patients that were randomized and received the assigned intervention.

#### 11.3 Primary study parameter(s)

A multivariable logistic regression model will be used to estimate the effect of convalescent neutralizing antibody containing treatment on disease severity. The following factors will be included in this analysis; sex, age at admission, CRP, oxygen saturation at room air, time since symptom onset of COVID-19 and COVID-19 vaccination status. A 2-sided Wald test on the OR of the treatment arm and the control arm based on the multivariable covariate-adjusted logistic regression analysis will be used to assess whether convalescent neutralizing antibody containing treatment reduces the incidence of a severe course of disease at a global alpha level of 5% and an adjusted alpha for one interim analysis of 4.82%.

#### 11.4 Secondary study parameter(s)

This analysis will describe the number patients who died after treatment, overall survival calculated from the date of randomization, including Kaplan-Meier curves per randomization arm, the duration of COVID-19 syndrome, the rate of viral clearance, and the development of long-term persistent neutralizing antibodies against SARS-CoV2. Also, the T-cell immunity as measured by in vitro specific T-cell response to COVID-19 tetrameric antigens will be analysed.

The Statistical Analysis Plan will be finalized prior to database lock.

#### 11.5 Other study parameters

Age and comorbidity (diabetes, obesity, cardiovascular risk) are known risk factors that segregate patients that have a high risk for a severe disease course.

##### 11.6 Interim analysis (if applicable)

In order to monitor safety and efficacy, two interim safety and one interim efficacy analysis will be done. A statistician with access to unblended data will report the results of both interim analyses to the DSMB. Based on the O'Brien Flemming alpha-spending function to account for one interim analysis, a *P*-value <0.0054 at the time of the interim efficacy analysis will be considered sufficient evidence to stop the trial. At the time of the interim analysis, the DSMB can advise the study team to increase the sample size of the study. Review of safety data (AEs, SAEs, SUSARs and laboratory outcomes) will be performed to check whether this gives the expected information on safety data of the study product.

| The DSMB will be asked to evaluate the safety and efficacy of the treatment and complications and provide advice on whether revision of the procedures is necessary. **Time point** | | **Analysis** |
|---|---|---|
| Interim-safety-analysis | First 10 randomized patients, all reached 28 days post last dosing | Safety:<br>All AEs of clinical interest and SAE, and Laboratory results, anti-COVID-19 antibody levels upon entry. |
| Interim-efficacy-analysis | First 43 randomized patients, all reached 28 days post last dosing | Safety and efficacy: anti-COVID-19 antibody levels upon entry. |
| Primary Analysis | 86 randomized patients | Safety and efficacy:<br>All AEs of clinical interest and SAE, and Laboratory results |

### 12. ETHICAL CONSIDERATIONS

#### 12.1 Regulation statement

The study will be conducted in accordance with the ethical principles of the Declaration of Helsinki, the ICH-GCP Guidelines, the EU Clinical Trial Directive (2001/20/EG), and applicable regulatory requirements. The site principal investigator is responsible for the proper conduct of the study at the study site. An accredited Ethics Committee will approve the study protocol and any substantial amendment.

#### 12.2 Recruitment and consent

Written informed consent of patients is required before enrolment in the trial and before any study related procedure takes place. The investigator will follow ICH-GCP and other applicable regulations in informing the patient and obtaining consent for the trial and for biobanking. The investigator should take into consideration if the patient is capable of giving informed consent. Before informed consent may be obtained, the investigator should provide the patient ample time and opportunity to inquire about details of the trial and to decide whether or not to participate in the trial. All questions about the trial should be answered to the satisfaction of the patient. There is no set time limit for the patient to make a decision. The investigator should inform each patient if there is a specific reason why he/she must decide within a limited time frame, for example if patients condition necessitates start of treatment or if the trial is scheduled to close for enrolment. The content of the patient information letter, informed consent form and any other written information to be provided to patients will be in compliance with ICH-GCP, GDPR and other applicable regulations and should be approved by the Ethics Committee in advance of use. The patient information letter, informed consent form and any other written information to be provided to patients will be revised whenever important new information becomes available that may be relevant to the patient’s consent. Any substantially revised informed consent form and written information should be approved by the Ethics Committee in advance of use. The patient should be informed in a timely manner if new information becomes available that might be relevant to the patient’s willingness to continue participation in the trial. The communication of this information should be documented.

#### 12.3 Benefits and risks assessment, group relatedness

COVID-19 is a syndrome with significant morbidity and mortality and patients with reduced B-cell function are at risk for more severe or more protracted disease courses. Adjunctive therapy with antibodies that are present in Nanogam that contain a certified amount of convalescent antibodies may be of aid in preventing a severe course of this disease or in shortening the syndrome duration. Although positive effects on these parameters of treatment with convalescent antibody containing therapy has not been shown to date, recently, a beneficial effect of convalescent plasma was proven in a cohort of elderly patients who were treated in first three days of disease (Libster, Perez Marc et al. 2021). Lack of effect of these therapies in later stages of disease may be a result of already present neutralizing antibodies in included patients, but whether convalescent antibody containing therapy is of aid in patients who do not develop neutralizing antibodies is currently unknown. Risks of convalescent antibody containing Nanogam plus are similar to those of conventional Nanogam and may consist of allergic reactions. Nanogam treatment in this population itself is widespread. Moreover, currently, compassionate use programs for treatment with convalescent plasma already exist for patients who are immunocompromised and treatment with these agents is performed frequently, although no solid evidence in terms of efficacy (such as trial results) exists. Taken together, the risk of side effects is small and treatment with convalescent plasma occurs outside of study protocols. Disease severity of COVID-19 may be high and the cohort of patients that may be included in this study will likely have no or limited benefit from vaccinations, as these patients are not likely to mount an effective protective immune response. We therefore deem the results of this study required to best aid the study population in courses of COVID-19 disease.

##### 12.4 Compensation for injury

The sponsor/investigator has a liability insurance which is in accordance with article 7 of the WMO.

The sponsor (also) has an insurance which is in accordance with the legal requirements in the Netherlands (Article 7 WMO). This insurance provides cover for damage to research subjects through injury or death caused by the study.

The insurance applies to the damage that becomes apparent during the study or within 4 years after the end of the study.

### 13. ADMINISTRATIVE ASPECTS, MONITORING AND PUBLICATION

Data management will be implemented according to Good Clinical Practice (GCP)- guidelines and supported by the independent Clinical Research Unit (CRU) of the AMC. The acquired imaging data will be stored on a file server in a directory only accessible to the investigators or system administrators. The executive investigator will construct and manage a database based on the study parameters (amongst which are the imaging data). A backup server is used for daily backups.

#### 13.1 Handling and storage of data and documents

Each patient is assigned a unique patient study number at enrolment. In trial documents the patient's identity is coded by patient study number as assigned at enrolment. The site principal investigator will keep a subject enrolment and identification log that contains the key to the code, i.e. a record of the personal identification data linked to each patient study number. This record is filed at the investigational site and should only be accessed by the investigator and the supporting hospital staff, and by representatives of the sponsor or a regulatory agency for the purpose of monitoring visits or audits and inspections. Patients confidentiality will be ensured in compliance with EU regulation and the Dutch Act on Implementation of the General Data Protection Regulation. (in Dutch: Uitvoeringswet AVG, UAVG).

#### 13.2 Handling and storage of data and documents

Data and documents will be controlled and processed conform the European Union (EU) General Data Protection Regulation (GDPR) and the Dutch Act on Implementation of the General Data Protection Regulation. (in Dutch: Uitvoeringswet AVG, UAVG).

#### 13.3 Filing of essential documents

Essential Documents are those documents that permit evaluation of the conduct of a trial and the quality of the data produced. The essential documents may be subject to, and should be available for, audit by the sponsor's auditor and inspection by the regulatory authority(ies).
The investigator should file all essential documents relevant to the conduct of the trial on site. The sponsor will file all essential documents relevant to the overall conduct of the trial. Essential documents should be filed in such a manner that they are protected from accidental loss and can be easily retrieved for review.

##### 13.4 Record retention

Essential documents should be retained for 25 years after the end of the trial. They should be destroyed after this time, unless a longer record retention period is required by site specific regulations.
Source documents (i.e. medical records) of patients should be retained for at least 15 years after the end of the trial described in section 18.4. Record retention and destruction after this time is subject to the site’s guidelines regarding medical records.

##### 13.5 Storage of data

Electronic patient data collected in the e-CRF will be stored at AmsterdamUMC for 25 years. Encoded SAE line listings will be shared with the manufacturer of the IMP for purpose of safety evaluation.

##### 13.6 Storage of samples

Biological samples should only be stored for the purpose of additional research if the patient has given consent. If no informed consent was obtained, samples should be destroyed after the patient has completed all protocol treatment and procedures. Storage of biological samples on site is subject to the site’s guidelines. Samples that are shipped to another facility (e.g. a central laboratory) for a purpose as described in this protocol or for additional scientific research, should be stripped from any identifying information and labeled with a code (trial name or number and patient study number as assigned at enrolment). Biological samples will be stored for a maximum of 5 years.

##### 13.7 Monitoring and Quality Assurance

Monitoring will be performed by the Clinical Research Unit (CRU) of the AmsterdamUMC location AMC. Appropriate monitoring and auditing procedures will be followed. The CRU will monitor the site for compliance with GCP guidelines, Dutch regulations, and Investigator obligations. The monitoring visits are also to provide the investigator with the opportunity to evaluate study progress; verify the accuracy and completeness of source data and CRF; and ensure that all protocol standards are being fulfilled. Monitoring visits may be conducted pre-study, during the study, and following study completion.

#### 13.8 Amendments

A ‘substantial amendment’ is defined as an amendment to the terms of the METC application, or to the protocol or any other supporting documentation, that is likely to affect to a significant degree:

- the safety or physical or mental integrity of the subjects of the trial;
- the scientific value of the trial;
- the conduct or management of the trial; or
- the quality or safety of any intervention used in the trial.

All substantial amendments will be notified to the METC and to the competent authority.

Non-substantial amendments will not be notified to the accredited METC and the competent authority, but will be recorded and filed by the sponsor.

#### 13.9 Annual progress report

The sponsor/investigator will submit a summary of the progress of the trial to the accredited METC once a year. Information will be provided on the date of inclusion of the first subject, numbers of subjects included and numbers of subjects that have completed the trial, serious adverse events/ serious adverse reactions, other problems, and amendments.

#### 13.10 Temporary halt and (prematurely) end of study report

The sponsor will notify the accredited METC and the competent authority of the end of the study within a period of 90 days. The end of the study is defined as the last patient’s last visit.

The sponsor will notify the METC immediately of a temporary halt of the study, including the reason of such an action.

In case the study is ended prematurely, the sponsor will notify the accredited METC and the competent authority within 15 days, including the reasons for the premature termination.

Within one year after the end of the study, the investigator/sponsor will submit a final study report with the results of the study, including any publications/abstracts of the study, to the accredited METC and the Competent Authority.

##### 13.11 Public disclosure and publication policy

Trial results will always be submitted for publication in a peer reviewed scientific journal regardless of the outcome of the trial – unless the trial was terminated prematurely and did not yield sufficient data for a publication.

All and any publications of (interim) trial results are subject to the AmsterdamUMC Publication Policy, according to the version of this policy that is effective at the time of publication. Public disclosure and publication policy is described in the Clinical Trial Agreement that has been signed for this study.

### 14. STRUCTURED RISK ANALYSIS

#### 14.1 Synthesis

Patients may have direct benefit from the treatment with Nanogam plus. No safety concerns are expected since these products have been registered and safety concerns are described in the SMPC. Immunoglobulins are normal components of the human body. Therefore, the usual preclinical toxicity studies in animals are not feasible due to overload of the circulation in acute toxicity tests and induction of antibodies in repeated dose studies.During the current study the patients will be intensively monitored and tolerance of Nanogam plus is assessed by measuring blood pressure, heart rate, and temperature before and directly after the infusion. Adverse events are monitored throughout the clinical study.

#### 16. Appendix 1

*WHO ordinal Scale for clinical improvement:*

| **Patient state** | **Descriptor** | **Score** |
|---|---|---|
| ***Uninfected*** | No clinical or virological evidence of infection | 0 |
| ***Ambulatory*** | No limitations of activities | 1 |
| | Limitation of activities | 2 |
| ***Hospitalized*** ***Mild disease*** | Hospitalized, no oxygen therapy | 3 |
| | Oxygen by Mask or nasal prongs | 4 |
| ***Hospitalized*** ***Severe Disease*** | Non-invasive ventilation or high-flow oxygen | 5 |
| | Intubation and mechanical ventilation | 6 |
| | Ventilation + additional organ support – vasopressors, renal replacement therapy (which was not required prior to disease), extracorporeal membrane oxygenation. | 7 |
| ***Dead*** | Death | 8 |

#### 17. Appendix 2

SMPC Nanogam
