## Supplemental Figure 1 for "SARS-CoV-2 hyperimmune globulin for severely immunocompromised patients with COVID-19: a randomised, controlled, double-blind, phase 3 trial"

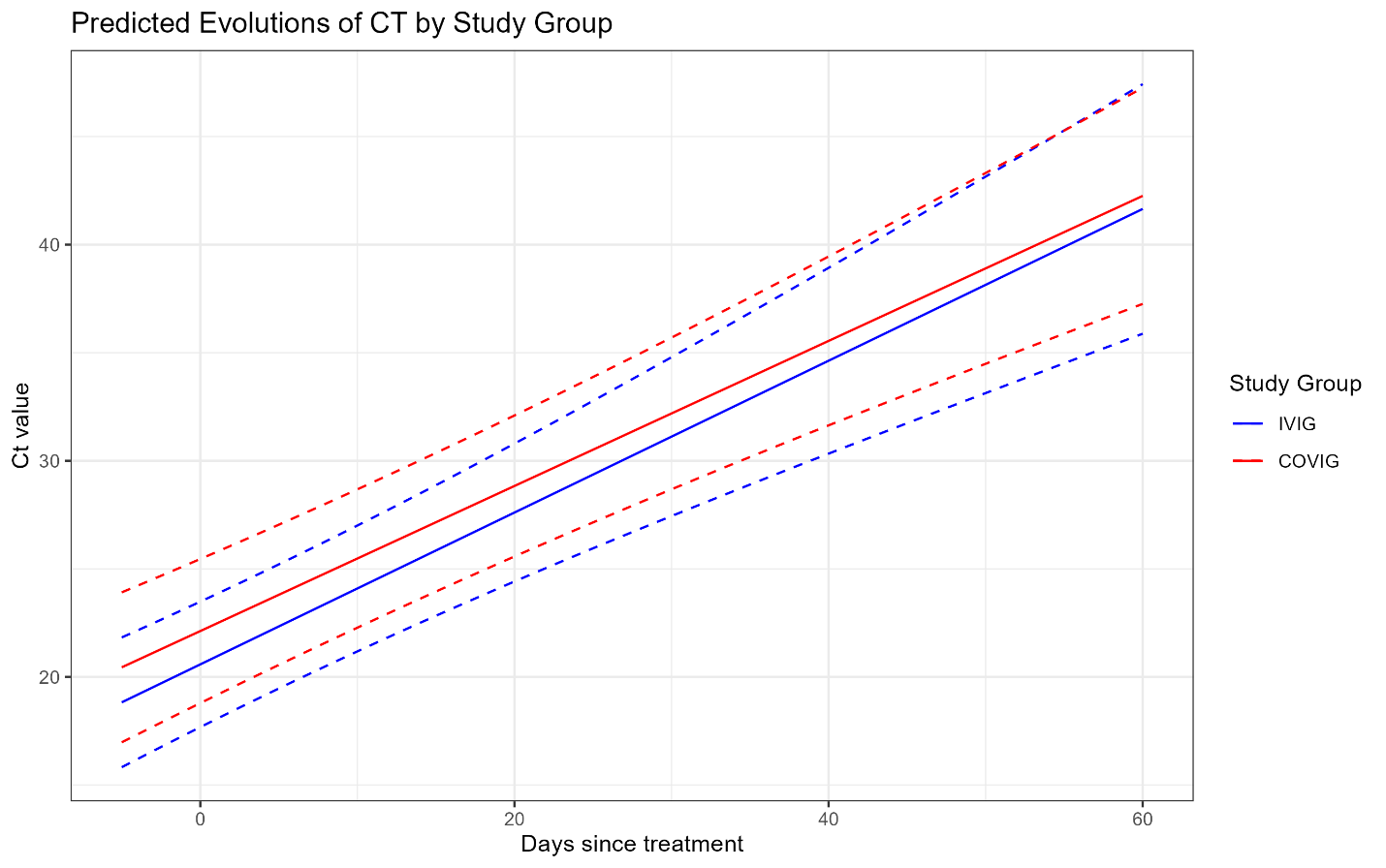

Figure 1: Predicted evolution of SARS-CoV-2 PCR Ct value by study group. The difference in the rate of change of PCR Ct value over time between the IVIG and COVIG groups was estimated at -0.016 (-0.141, 0.110, p = 0.802) meaning that there was no statistically significant difference in the rate of change of Ct value between the two groups.
